# Addressing Cervical Cancer Screening Through Self-Sampling and HPV Testing Among Under-Screened Women: A Case Study in the Decentralized Portuguese Setting

**DOI:** 10.1101/2024.04.05.24305323

**Authors:** Sara da Graça Pereira, Luís Nobre, Marina Ribeiro, Patrícia Carvalho, Ana Morais, Rita Sousa, Ana Paula Moniz, Francisco Matos, Graça Fernandes, João Pedro Pimentel, José Carlos Marinho, José Luís e Sá, Olga Ilhéu, Teresa Rebelo, José Fonseca-Moutinho, Hugo Prazeres, Rui Jorge Nobre, Fernanda Loureiro

## Abstract

**Background:** Cervical cancer (CC) screening is crucial for reducing its incidence. However, encouraging participation among under-screened women remains challenging. Portugal’s decentralized health regions provide an ideal case study due to a significant proportion of eligible women avoiding regular screening. Globally, self-sampling has emerged as a promising solution to enhance screening attendance. This study aims to assess self-sampling acceptance among under-screened women in central Portugal, contributing to the existing knowledge of self-sampling in CC screening.

**Methods:** 801 women aged 30-59, not participating in the Central Region’s CC Screening for 4 or more years, were randomly recruited. Women who accepted to participate in the study received cervicovaginal self-sampling kits at home. Women with a positive high-risk human papillomavirus (hr-HPV) test result were invited for gynaecological follow-up.

**Results:** Among the 687 eligible women, 307 (44.7%) accepted, and 198 (28.8%) provided specimens for hr-HPV testing. Out of twelve positive cases, eleven underwent gynaecological follow-up, identifying six cervical lesions.

**Conclusions:** The study highlights the potential of self-sampling and HPV testing to enhance CC screening in Portugal, with encouraging acceptance and effective detection of cervical lesions. These findings offer a promising solution for addressing under-screening among eligible women in the decentralized health regions of Portugal.

## Introduction

Cervical cancer (CC) remains a significant global health concern, despite established screening programmes, vaccination, and treatment options. As the fourth most common and fatal cancer among women worldwide^1^, CC claims the lives of over 300,000 women annually, equivalent to one death every two minutes. Portugal, among Southern European countries, reported the fourth-highest age-standardized CC incidence rate in 2020, reaching 10.7 incidences and 3.2 mortalities per 100,000 women according to the International Agency for Research on Cancer (IARC)^2^.

Organized CC screening programmes using cervical cytology have made progress in reducing global incidence and mortality rates. However, challenges such as the low sensitivity and weak inter-rater reproducibility of cytology^3, 4^, a shortage of general practitioners, logistical difficulties in scheduling appointments, and psychological barriers (including past clinical visit pain, discomfort with genital exposure, history of sexual abuse, intimate partner violence, religious/cultural beliefs, as well as limited knowledge about the disease^5-7^) impede its effective implementation. These persistent barriers restrict some women’s access to conventional CC screening programmes.

Several studies reveal that most new CC cases are identified in women with infrequent or no screening history, and, in some of cases, the malignancy emerges even in women with consistently negative cytological results before malignancy is diagnosed^8-10^. This often leads to later-stage diagnoses, more invasive treatments, and reduced quality of life for these women. Hence, there is an urgent need for innovative screening methods to address these challenges.

HPV is recognized as the primary cause of CC, responsible for nearly all cases^11-13^. Numerous studies consistently demonstrate that HPV testing in CC screening surpasses cytology in accurately detecting pre-cancerous cervical lesions, making it more effective in reducing disease incidence and mortality^14-16^. This method is highly reproducible and reliable and allows for longer screening intervals, especially for women who test negative for HPV. This not only eases the burden on healthcare systems but also enhances overall screening efficiency^17^. Recognizing these advantages, the World Health Organization (WHO) now recommends the HPV test as the primary strategy for CC screening^18^.

The Portuguese Society of Gynaecology recommends regular CC screening for all women aged 25 to 60 years^19^. At the date of this study, cervical cytology triages every three years with HPV testing for Atypical Squamous Cells of Undetermined Significance (ASC-US) cases was the protocol in force in Portugal. Nowadays, the recommended screening algorithm is the search for oncogenic HPV genotypes in cervicovaginal samples every five years. The implementation of this strategy is currently underway in several regions of Portugal^19, 20^.

Despite efforts to implement organized CC screening programmes regionally, coverage rates in Portugal fell below expectations. At the time of our study, in 2019^21^, while geographical coverage in Portugal reached 98.4%, the annual population coverage was only 52.8%. This significant gap suggests that about half of eligible women did not receive a call for screening. Moreover, suboptimal adherence rates (76%) highlight significant opportunities for improvement.

A recent study highlighted factors influencing the low participation of Portuguese women in CC screenings, including doctor unavailability, work schedule conflicts, disease ignorance, and discomfort with male doctors, among others^22^. Another study noted lower screening adherence among economically disadvantaged and less health-conscious Portuguese women^23^. For migrant women in Portugal, non-attendance is associated with younger age, being born in Africa or Asia, single/divorced/widowed status, no previous consultation with a General Practitioner in Portugal, and infrequent gynaecological check-ups^24^.

The adoption of the HPV test as the primary method for CC screening has facilitated the introduction of self-sampling tests, allowing women to collect their own samples. This method overcomes critical barriers to CC screening, providing a convenient, private, and emotionally comfortable solution, potentially increasing screening attendance and reaching women who would otherwise not undergo screening^25^.

Recent worldwide research consistently demonstrates that offering self-sampling kits for hr-HPV testing significantly increases screening numbers, particularly among women not reached by in-clinic programmes^5, 26-30^. A comprehensive meta-analysis comprising 154 HPV self-sampling studies worldwide, involving a total of 482,271 women, concluded that self-sampling procedures had a substantial impact on the likelihood of CC screening uptake, nearly doubling the probability (RR: 1.8; 95% CI: 1.7–2.0) compared to clinician-collected samples^31^.

Some metanalysis have shown that self-sampling tests, particularly when coupled with hr-HPV testing using PCR technology, exhibit comparable sensitivity to detect cervical intraepithelial neoplasia (CIN) 2+ (CIN2, CIN3, or cancer) when compared to samples collected by healthcare professionals^32, 33^. These findings provide strong evidence supporting the suitability of self-collected samples for CC screening.

Our study represents the pioneering effort to **assess self-sampling acceptance**, combined with the **hr-HPV test, to encourage women who are not regular participants in the organized CC screening programme within Portugal’s central region**.

The implementation of self-sampling for CC screening in Portugal shows great potential, thanks to a well-established primary care network that serves as a robust foundation for distributing kits and providing guidance on usage. The flexibility allowed by Regional Health Administrations enables personalized management, considering the specificities of each region, facilitating effective coordination of logistics healthcare professional training, and public education. Moreover, this decentralized framework encourages direct engagement and collaboration with local communities, fostering acceptance and adherence to self-sampling. Our investigation also highlights the effectiveness of various workflow elements, such as reminder phone calls and letters. Additionally, it explores the impact of providing self-sampling kits exclusively to consenting participants rather than distributing them to all eligible women. These findings are invaluable for enhancing the efficiency of CC screening.

## Material and Methods

### Study Design and Sample Selection

The present study received approval from the Ethics Committee of the Central Regional Health Administration (Study nº 23/2018), and was conducted from October 2018 to September 2019, predating the COVID-19 pandemic in the central region of Portugal.

A total of 801 women were randomly selected from the CC screening database of the Central Region of Portugal. To be included in the study, women had to meet four specific criteria: i) aged 30-59 years, ii) with no cytology results in the past four years, iii) not pregnant, and iv) without a recorded hysterectomy. Randomization was stratified based on six different health centre groups (HCG): ‘Baixo Mondego’, ‘Baixo Vouga’, ‘Dão-Lafões’, ‘Pinhal Interior Norte’, ‘Pinhal Litoral’ and ‘Castelo Branco Local Health Unit’ (**Table 1**). Stratification aimed to ensure representation of each HCG in the study population. This selection was based on available records within these groups, providing accurate data on women who had not participated in the screening programme for four or more years. Furthermore, stratification occurred across three age groups (30-39, 40-49, and 50-59 years) to examine potential age-related influences on women’s participation.

**Table 1.** Characterization of the Study Population: Number of Selected Women in Each Age Group and for Health Centre Group.

| Age (years) | Health Centre Groups |  |  |  |  |  | Total |
| --- | --- | --- | --- | --- | --- | --- | --- |
|  | Baixo Mondego | Baixo Vouga | Dão-Lafões | Pinhal Interior Norte | Pinhal Litoral | LHU Castelo Branco |  |
| 30-39 | 69 | 62 | 55 | 25 | 50 | 6 | 267 |
| 40-49 | 70 | 60 | 54 | 26 | 52 | 5 | 267 |
| 50-59 | 66 | 60 | 53 | 30 | 52 | 6 | 267 |
| Total | 205 | 182 | 162 | 81 | 154 | 17 | 801 |

### Invitation letter sending, self-sampling kit shipping, reminder phone call A and B and final reminder letter

The study flowchart is illustrated in **Figure 1**. At the study’s onset (week 0), all selected women received an invitation letter outlining the study and an informed consent form. This form allowed them to confirm their interest in participating and request a self-sampling kit for cervicovaginal fluid collection, intended for subsequent hr-HPV testing.

**Figure 1.**
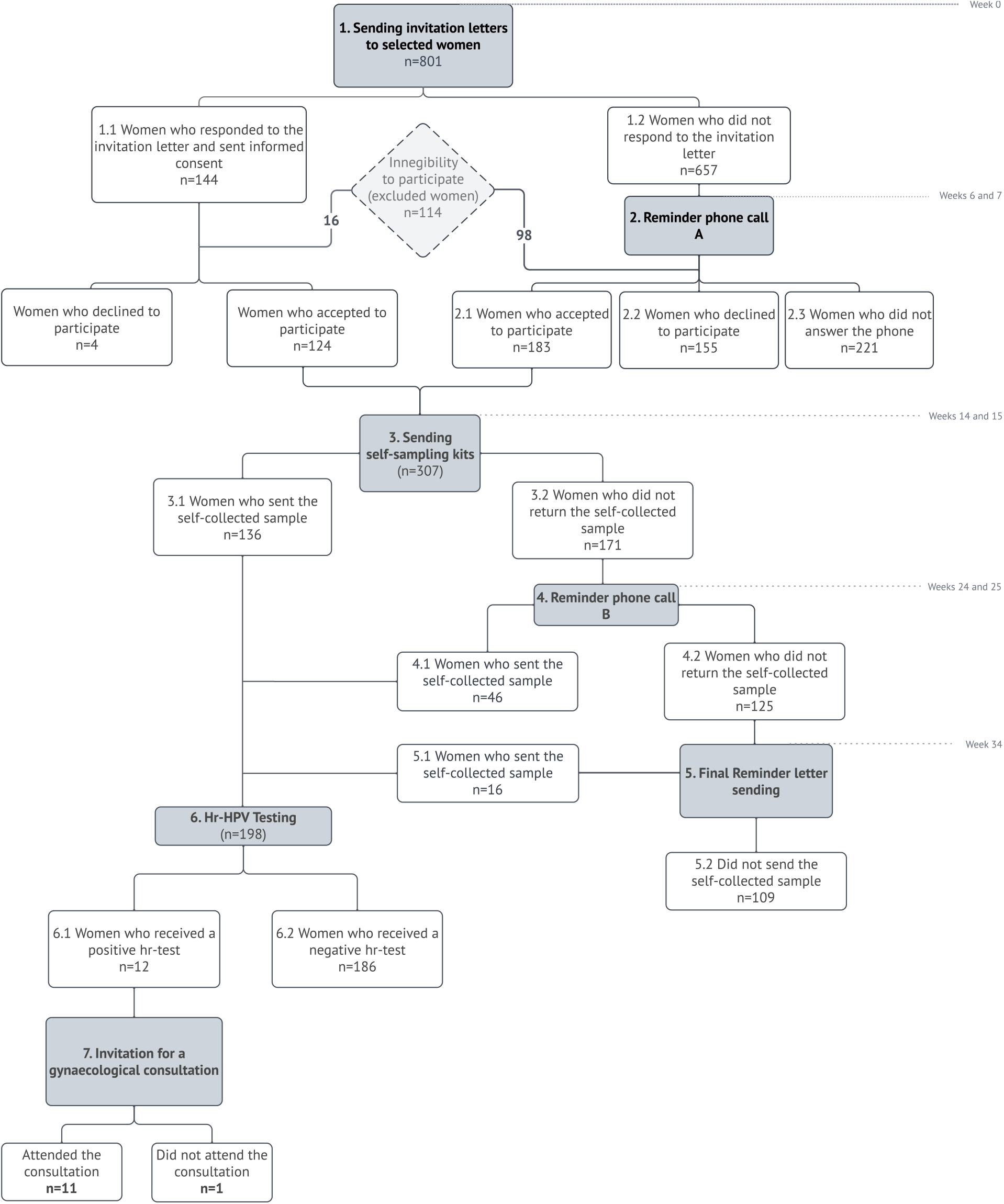
Study Flowchart: Involvement of Participants Throughout Various Phases of the Study. This flowchart outlines the number of women who entered each phase of the study. Women were excluded from the study for various reasons, including health conditions that impeded sample collection, absence from the country, death, pregnancy, hysterectomy, recent participation in the regional CC screening programme, or inability to reach women, due to unsuccessful attempts to establish contact.

Women who did not initially respond received a follow-up phone call (reminder phone call A) six weeks after the invitation to confirming the receipt of the invitation letter (weeks 6 and 7 of the study). Those expressing their interest in participating, either by signing the informed consent form or during reminder phone call A, received a self-sampling kit during weeks 14 and 15 of the study. Each kit included a Qvintip sampler (*Aprovix AB, Solna, Sweden*), a labelled test tube with a cap, a data form, an instruction manual for self-sampling, storage, and shipment of the biological material, as well as a prepaid and pre-addressed envelope for sample shipment. Ten weeks after the kit distribution (weeks 24 and 25), women who had not yet sent the sample were contacted by phone for sample return -reminder phone call B. Finally, at week 34, a final reminder letter was dispatched to women who had not returned the biological sample, informing them of the study’s closure at week 41, after which no more samples would be accepted.

### Nucleic acid extraction and hr-HPV testing in self-collected samples

DNA was extracted from self-collected samples using the *RealLine DNA-Express kit – VBC8899-R*, (Bioron Diagnostics GmbH, Römerberg, Germany). The presence of 12 hr-HPV types was detected by Real-Time PCR (qPCR) using the *RealLine HPV HCR Genotype Fla-Format – VBD8482, a CE-IVD* commercial kit assay (Bioron Diagnostics GmbH) following the manufacturer’s instructions. This test detects and identifies the following hr-HPV genotypes individually: 16, 18, 31, 33, 35, 39, 45, 51, 52, 56, 58 and 59. Additionally, it evaluates the quality of the extracted DNA by amplifying the endogenous beta-actin gene. A report with the hr-HPV test result (positive or negative) was sent to each of the women who submitted the self-collected sample.

### Gynaecology consultation, Cytological diagnosis, hr-HPV testing and Anatomopathological examination

Women who tested positive for hr-HPV infection were referred to the colposcopy unit at the Portuguese Institute of Oncology in Coimbra three months after receiving their results. At their first visit, they repeated self-sampling using the Qvintip kit and underwent a gynaecological exam, including clinical evaluation, cervicovaginal sampling for co-testing with cytology and HPV, and colposcopy with biopsy if any abnormalities were found.

The detection and genotyping of hr-HPV infection from self-collected samples were conducted using the methodology outlined previously. The detection of hr-HPV infection in cytological samples was performed with clinically validated Cobas® HPV test, which detects 14 hr-HPV types: HPV 16, HPV 18 and other hr-HPV types (31, 33, 35, 39, 45, 51, 52, 56, 58, 59, 66, and 68). Pap-test cytology was conducted by certified pathologists following the Bethesda classification system.

These procedures followed established best practices at the Gynaecological and Pathological Anatomy Laboratories of the Portuguese Institute of Oncology. The results were used to categorize diagnoses according to the CIN classification system.

## Results

### Women’s Participation and the Impact of Reminder Strategies in the Study

The study aimed to assess the feasibility of self-sampling combined with HPV testing as an additional method to conventional CC screening for women not regularly engaged in it. A total of 801 Portuguese women from the central region, who had not undergone screening for four years or more, were invited to participate through an invitation letter.

Of the 801 women invited, 144 returned the informed consent form included in the invitation letter with the majority doing so within the first two weeks. Of these, 124 agreed to participate in the study, 4 declined, and 16 did not meet the eligibility criteria. The remaining 657 women did not respond in the first 5 weeks of the study and were, therefore, contacted via phone call (reminder phone call A) in weeks 6 and 7 of the study. From this group, 183 women agreed to participate, 155 declined, 221 did not answer the phone and 98 did not meet the eligibility criteria.

In total, 114 women (14.2%) were excluded from the study. This included those who did not meet criteria such as recent participation in the CC screening programme, hysterectomy, and pregnancy. Exclusions also resulted from unsuccessful efforts to make contact, such as returned letters and unsuccessful phone calls due to incomplete address and contact details.

Additionally, exclusions were made for other reasons, such as health conditions impeding sample collection, death, or absence from the country (**Table S1**). Ultimately, 307 out of 687 women (44.7%) agreed to participate in the study and requested the self-sampling kit. Among them, 198 women collected and sent cervicovaginal fluid samples for subsequent hr-HPV testing. Out of these samples, 136 were received within the initial 11 weeks after the distribution of self-sampling kits (in weeks 14 and 15 of the study). Additionally, 46 samples were only received following reminder phone call B at week 25 of the study. Lastly, an additional 16 samples were received upon sending the final reminder letter at week 34 of the study (**Figure S1**).

Analysing the study’s population participation, data showed a slightly higher involvement rate among women in the ‘50-59 age group’ (32.4%), followed by women in the ‘40-49 age group’ (29.2%) and women in the ‘30-39 age group’ (25.1%) (**Table 2**). In terms of various Health Care Groups (HCG), the highest participation rate was observed in the Dão-Lafões HCG (31.9%), followed by Baixo Mondego (30.7%), Baixo Vouga (28.3%), Pinhal Litoral (27.7%), and Pinhal Interior Norte (26.8%). No women from the ULS de Castelo Branco participated in the study (**Table 2**).

**Table 2.** Participation Rate and hr-HPV Prevalence Across Different Health Centres and Age Groups Considered.

| Age Group | Total |  |  | 30-39 |  |  | 40-49 |  |  | 50-59 |  |  |
| --- | --- | --- | --- | --- | --- | --- | --- | --- | --- | --- | --- | --- |
| Health Centre Group | Eligible Women Invited (n) | Participant n (%) | Positive hr-HPV Samples n (%) | Eligible Women Invited (n) | Participant n (%) | Positive hr-HPV Samples n (%) | Eligible Women Invited (n) | Participant n (%) | Positive hr-HPV Samples n (%) | Eligible Women Invited (n) | Participant n (%) | Positive hr-HPV Samples n (%) |
| Baixo Mondego | 179 | 55(30.7) | 2(3.63) | 62 | 15(24.2) | 1(6.7) | 58 | 20(34.5) | 1(5.0) | 59 | 20(33.9) | 0(0.0) |
| Baixo Vouga | 145 | 41(28.3) | 2(4.9) | 53 | 14(26.4) | 1(7.1) | 47 | 16(34.0) | 1(6.3) | 45 | 11(24.4) | 0(0.0) |
| Dão-Lafões | 141 | 45(31.9) | 2(4.44) | 49 | 15(30.6) | 2(13.3) | 48 | 13(27.1) | 0(0.0) | 44 | 17(38.6) | 0(0.0) |
| Pinhal Interior Norte | 71 | 19(26.8) | 1(5.26) | 22 | 6(27.3) | 1(16.6) | 22 | 6(27.3) | 0(0.0) | 27 | 7(25.9) | 0(0.0) |
| Pinhal Litoral | 137 | 38(27.7) | 5(13.1) | 48 | 10(20.8) | 3(30.0) | 48 | 11(22.9) | 0(0.0) | 41 | 17(41.5) | 2(11.8) |
| Castelo Branco LHU | 14 | 0(0.0) | 0(0.0) | 5 | 0(0.0) | 0(0.0) | 3 | 0(0.0) | 0(0.0) | 6 | 0(0.0) | 0(0.0) |
| All | 687 | 198(28.8) | 12(6.1) | 239 | 60(25.1) | 8(13.3) | 226 | 66(29.2) | 2(3.0) | 222 | 72(32.4) | 2(2.8) |

### HPV Analysis Results and Cytological-Histological Evaluation of Positive Cases

Hr-HPV types were detected in 12 out of the 198 samples (6.1%). The incidence of hr-HPV infection was 13.3% for women in the 30–39 age group, 3% for women in the 40-49 age group and 2.8% for women in the 50-59 age group, indicating a decrease in hr-HPV infection with age (**Table 2**).

Women who tested positive for hr-HPV were referred to the colposcopy unit, with 11 out of 12 women (92%) attending. The results of HPV tests from self-sampling procedures, cytology, colposcopic findings, and histological analysis, are outlined in **Table 3**.

**Table 3.** Correspondence of hr-HPV Test Results in Self-collected Cervicovaginal Fluid Samples and in Cytological samples and Their Correlation with Cytological, Colposcopic, and Histological Analyses.

| Age group | Woman ID | 1st hr-HPV test (self-sampling) | 2nd hr-HPV test (self-sampling) | Cytology | Colposcopy | Histology |
| --- | --- | --- | --- | --- | --- | --- |
| 30-39 | 64 | HPV58 | Negative | NILM | Normal | NP |
|  | 237 | HPV45 | HPV45 | ASC-US | Abnormal colposcopic findings (grade 1) | LSIL/CIN1 |
|  | 397 | HPV51 | HPV51 | ASC-US | Abnormal colposcopic findings (grade 1) | LSIL/CIN1 |
|  | 439 | HPV16 | HPV16 | NILM | Abnormal colposcopic findings (grade 2) | LSIL/CIN1 |
|  | 571 | HPV45 | HPV45 | ASC-US | Abnormal colposcopic findings (grade 2) | Inflammatory alterations |
|  | 644 | HPV31, HPV51 | HPV31, HPV51 | ASC-US | Abnormal colposcopic findings (grade 1) | LSIL/CIN1 |
|  | 674 | HPV31, HPV33 | HPV31, HPV33 | ASC-H | Abnormal colposcopic findings (grade 1) | HSIL/CIN2 |
| 40-49 | 138 | HPV31 | HPV31 | NILM | Normal | NP |
|  | 308 | HPV16 | HPV16 | ASC-US | Abnormal colposcopic findings (grade 2) | LSIL/CIN1 |
| 50-59 | 749 | HPV51 | HPV51 | ASC-US | Normal | NP |
|  | 754 | HPV39 | HPV39 | NILM | Normal | NP |

HPV – Human Papilloma Virus; NILM – Negative for Intraepithelial Lesion or Malignancy; ASC-US – Atypical Squamous Cells of Undetermined Significance; ASC-H – Atypical Squamous Cells, Cannot Exclude a High-Grade Squamous Intraepithelial Lesion; NP – Not Performed; LSIL – Low-Grade Squamous Intraepithelial Lesion; CIN1 – Cervical Intraepithelial Neoplasia Grade 1; HSIL – High-Grade Squamous Intraepithelial Lesion; CIN2 – Cervical Intraepithelial Neoplasia Grade 2.

Regarding the HPV tests conducted on self-sampling samples, there was nearly complete agreement (91%) between the results obtained from the first and second self-collected samples, except for one case (ID64), which turned negative.

The histological analysis revealed one case of non-neoplastic change (inflammatory alteration), five cases of CIN1 (mild dysplasia), and one case of CIN2 (moderate dysplasia). In all these women, an hr-HPV infection was detected in the corresponding self-collected sample.

## Discussion

Screening programmes play a pivotal role in curbing CC incidence and mortality globally^11, 34, 35^. Nevertheless, several countries, including Portugal, struggle with insufficient or non-existent CC screening coverage. In Portugal, only approximately 50% of eligible women undergo screening, leaving a significant percentage unscreened^21^.

Since a significant proportion of CC cases occur in under-screened or never screened women^32, 36^, it is important to encourage women to participate in screening programmes. Various global studies^37, 38^, supported by the WHO, have demonstrated that self-sampling significantly increases the involvement of women who are usually harder to reach in screening processes. This method enables women to collect their own samples privately and conveniently, overcoming obstacles that typically prevent their engagement in CC screening. Our study marks the first attempt to test HPV self-sampling for CC screening among non-screened women in Portugal. To achieve that, 801 women, between the ages of 30 and 59, were randomly selected from Central Portugal’s CC screening database following specific criteria.

Out of the 687 eligible women, we successfully obtained 198 cervicovaginal fluid samples through self-sampling, resulting in an overall attendance rate of 28.8%. These rates are consistent with previous studies in other populations, where participation in self-sampling groups ranged from 10.2% to 39%^5, 8, 26, 28, 29, 39-47^. Our data indicate also that self-sampling appears to be suitable for women of all ages covered by the Portuguese screening programme. Out of the 198 cervical fluid samples obtained, 72 samples belonged to the 50-59 age group (36.4%), 66 to the 40-49 age group (33.3%), and 60 to the 30-39 age group (30.3%). This finding is supported by several other studies^28, 29, 44, 47^.

Our study used an ‘opt-in’ strategy, where the self-sampling kit was sent only to women who expressed interest, rather than distributing it to all eligible women. The ‘send-to-all’ or ‘opt-out’ strategy, which involves delivering the self-sampling device to all enrolled women, has been shown to be more efficient in terms of participation rates, as suggested by three different meta-analyses studies^10, 31, 32^. In fact, this strategy allows women to familiarize themselves with the self-sampling device before making their decision. Therefore, implementing an ‘opt-out’ strategy in the future could be beneficial in boosting participation rates and reaching a larger number of hard-to-reach women.

Several other strategies which have proven to be successful in increasing women’s participation include invitations through community campaigns and door-to-door visits. These approaches have shown twice the participation rates compared to traditional invitation letters or reminders for standardized screenings^17, 32^.

Although we did not specifically include a questionnaire to understand why some women did not participate in our study, we gained insights during reminder phone calls from 159 non-participants. Most of them (58) mentioned having private health insurance, while a few cited scheduling or availability issues (5). Some expressed concerns about discomfort, pain, or doubts about correctly performing the self-sampling (6), while others doubted its effectiveness (4) or preferred the traditional cytological examination (4). The fear of not properly performing self-sampling seems to be a common concern, as concluded by Nishimura and co-workers in a systematic review encompassing 72 studies and 52,114 women^25^.

In future investigations, using a detailed questionnaire to address these concerns could be valuable. Such a questionnaire could help identify strategies to reach even more women and uncover other problems for which self-sampling may be a suitable solution.

Following the receipt and anonymization process of the samples, DNA was extracted and tested for quality and for the presence of hr-HPV infection. All 198 collected samples showed an acceptable amplification for the endogenous gene, indicating its suitability for hr-HPV analysis. This success may be credited to the user-friendly nature of the selected medical device (Qvintip®). According to Nishimura, cervical swabs, like the one used in our study, are preferred by women for self-sampling over other devices like lavage tools, cervical brushes, tampons, or labial pads^25^. Interestingly, some of the highest participation rates were observed using this device^8, 29^, and other reports have also highlighted its ease of use compared to other devices^49, 50^.

In our study, hr-HPV was detected in twelve women (6.1%). This may seem lower compared to other studies in Portugal. For instance, Rosário et al.^51^ found a 12.5% prevalence of hr-HPV in a CC screening study in the north of the country, and the CLEOPATRE study^52^ reported a 19.4% prevalence among women attending national gynaecology/obstetrics or STD clinics. However, it is important to consider our study’s population. In fact, we looked at 687 women, taking 198 cervical samples, which is relatively small. Also, our study focused on women aged 30 and older, while the other studies included younger women starting from 18 or 24 years old.

Still, our findings are consistent with a meta-analysis conducted by Arbyn and colleagues, which looked at twenty-two trials^32^, and found hr-HPV prevalence rates among under-screened or never-screened women ranging from 6.0% to 29.4% in self-sampling strategies.

Regarding the presence of hr-HPV infection by age group, there was a decrease in hr-HPV prevalence as women age, as documented in other studies^53, 54^. This observation aligns also with findings from other self-sampling studies^8, 42^.

In our study, out of the 12 women with an hr-HPV infection, 11 agreed to undergo a gynaecological appointment. This high acceptance rate for medical monitoring suggests that even though our target population might have underestimated or neglected screening, they are willing to seek medical attention upon receiving a positive result. This finding is consistent with other studies on self-sampling, underscoring the potential of self-sampling as a bridge between initial screening and necessary medical intervention^39, 42, 45, 47, 55^.

Meta-analyses across multiple studies involving screening and referral cohorts have consistently revealed that the sensitivity of HPV testing on self-collected samples, particularly when utilizing PCR-based HPV DNA tests, is statistically comparable to that observed with clinician-obtained cervical samples in detecting CIN2+^32, 33^. Although our sample size may limit the ability to draw definitive conclusions, it does suggest at least that self-sampling with HPV testing does not contradict conventional sampling methods. Interestingly, among hr-HPV positive women, a majority (7 out of 11) exhibited abnormal colposcopy findings with confirmed histological alterations. Importantly, our data underscore the significance of maintaining contact with patients and reminding them about the importance of screening. Reminder call B and final reminder letter resulted in an additional 62 cervicovaginal samples being submitted for analysis (around 31% of all specimens). This finding is consistent with other studies conducted in other countries^28, 29, 39^. Conversely, studies lacking reminder strategies in self-sampling for CC screening tend to have low participation rates^47^.

While our study provides valuable insights into the potential of self-sampling on CC screening rates in Portugal, it is essential to acknowledge certain limitations that may affect interpreting the results and their wider application. In fact, our participant group was relatively modest, starting with 801 women but narrowing down to 687. This reduced number may not holistically represent the diverse spectrum of women who either do not participate or participate irregularly in screenings in Portugal. Future studies could aim for a larger and more diverse participant pool to improve the findings’ applicability and understanding of under-screened women’s responses to self-sampling.

The absence of a control group called for conventional screening using cervical cytology limits the depth of comparative analysis. Including such a group would have allowed a more comprehensive assessment of self-sampling acceptance rates compared to traditional methods. Future research incorporating a control group undergoing conventional screening would provide a clearer understanding of self-sampling’s effectiveness relative to established approaches.

In summary, our study underscores the potential impact of integrating self-sampling with hr-HPV testing as a complementary approach to organized screening programmes. This strategy holds promise in improving CC screening rates, especially among less engaged women. This research’s importance is underscored by identifying hr-HPV infections that resulted in a notably high rate of subsequent gynaecological follow-up. Several cases exhibited abnormal colposcopic findings, which were later confirmed through histology.

The past three years have been heavily influenced by the COVID-19 pandemic, leading to global efforts to control the virus and ease strain on healthcare systems. This included widespread lockdowns, resource reassignment, and the postponement of non-urgent medical procedures, inadvertently disrupting preventive healthcare in Europe. HPV vaccination and CC screening were notably affected, raising concerns among experts about potential long-term impacts on CC rates and mortality due to these disruptions^56^. In response to these concerns, attention has shifted towards the effectiveness of self-sampling in detecting cervical lesions. The WHO has endorsed primary HPV-based screening and recently included self-sampling in guidelines for self-care interventions in health and well-being^57^. Furthermore, the IARC updated its assessment of CC screening methods, further supporting the recommendation for self-sampling^58^. Amidst challenges like the pandemic, self-care strategies, including self-sampling for HPV testing, emerge as crucial solutions. This approach not only addresses immediate concerns but also offers a sustainable solution for preventive healthcare.

In conclusion, our research demonstrates the effectiveness of self-sampling for HPV testing in improving participation rates in CC screening programs, especially among under-screened women in Portugal. Our findings also suggest that self-sampling is a valid approach, as evidenced by the agreement between the results of HPV testing conducted on self-collected samples and those obtained through gynaecological sampling. However, additional studies in a larger population are needed to validate these results. If confirmed, this method could receive official recognition as a complementary approach to current screening practices, thus aiding the overarching goal of conventional screening to lower the mortality rate associated with this cancer in the country.

## Data Availability

All data produced in the present work are contained in the manuscript

## Acknowledgements

The authors would like to express their gratitude to all the women who voluntarily participated in the present study. Additionally, appreciation goes to the entire technical staff of the Department of Public Health of the Central Regional Health Administration (ARS Centro, Portugal) and the Portuguese Institute of Oncology at Coimbra.

