## Supplemental data for "Addressing Cervical Cancer Screening Through Self-Sampling and HPV Testing Among Under-Screened Women: A Case Study in the Decentralized Portuguese Setting"

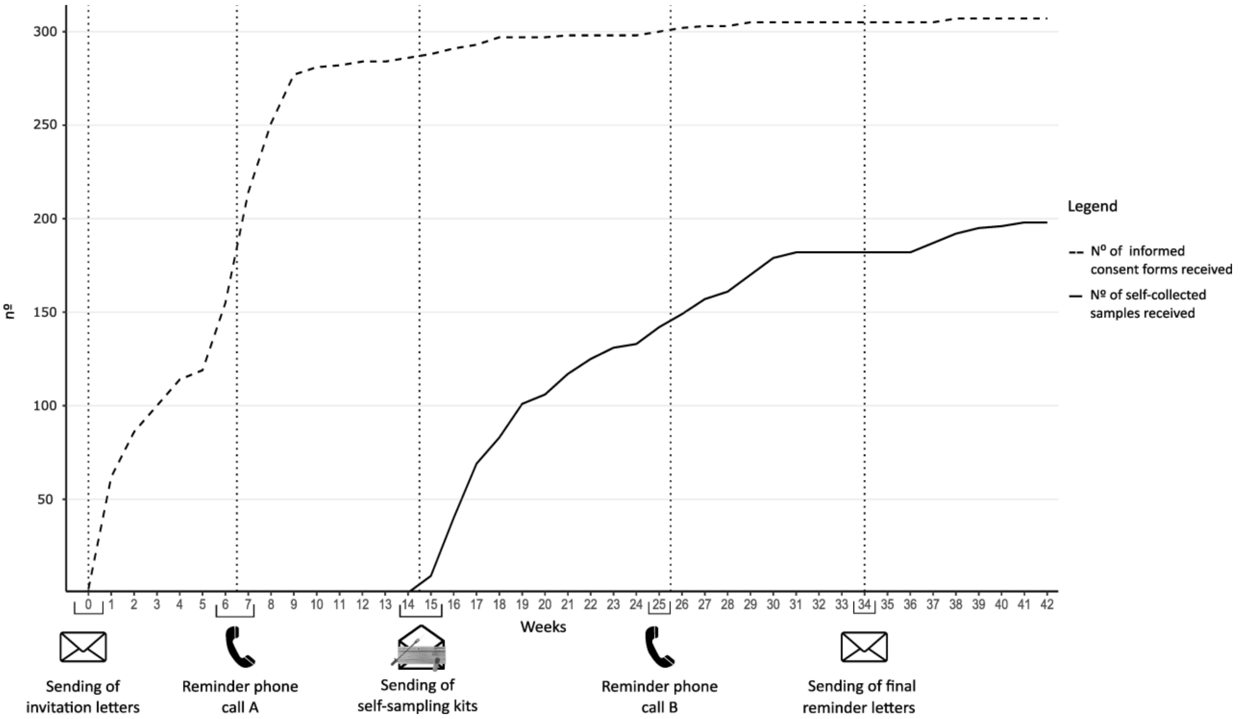


**Figure S1 – Correlation Between the Reception of Informed Consents, indicative of Participation Acceptance, and the Subsequent Return of Self-Collected Samples.**

**Table S1 – Reasons for Women's Ineligibility to Participate in the Study: Responses from the Invitation Letter and Reminder Phone Calls.**

| Factors Contributing to Women's Ineligibility for Participation in the Study | | n(%) |
|---|---|---|
| **Did not fulfil the study criteria** | | |
| | Recent participation in CC screening program | 43(37.7) |
| | Hysterectomised | 32(28.0) |
| | Pregnant | 5(4.4) |
| **Other Reasons** | | |
| | Unable to perform self-sampling due to underlying health conditions | 5(4.4) |
| | Deceased | 2(1.8) |
| | Absence from the country | 19(16.7) |
| **Insufficient address/contacts** | | 8(7) |
| **Total** | | 114(100) |
